# The Possible Role of Vitamin D in Suppressing Cytokine Storm and Associated Mortality in COVID-19 Patients

**DOI:** 10.1101/2020.04.08.20058578

**Authors:** Ali Daneshkhah, Vasundhara Agrawal, Adam Eshein, Hariharan Subramanian, Hemant K. Roy, Vadim Backman

## Abstract

**Objectives:** To investigate the possible role of Vitamin D (Vit D) deficiency via unregulated inflammation in COVID-19 complications and associated mortality.

**Design:** The time-adjusted case mortality ratio (T-CMR) was estimated as the number of deceased patients on day N divided by the number of confirmed cases on day N-8. The adaptive average of T-CMR (A-CMR) was further calculated as a metric of COVID-19 associated mortality in different countries. A model based on positivity change (PC) and an estimated prevalence of COVID-19 was developed to determine countries with similar screening strategies. Mean concentration of 25-hydroxyvitamin D (25(OH)D) in elderly individuals in countries with similar screening strategies were compared to investigate the potential impact of Vit D on A-CMR. We analyzed data showing a possible association between high C-Reactive Protein (CRP) concentration (CRP ≥ 1 mg/dL) and severe COVID-19. We estimated a link between Vit D status and high CRP in healthy subjects (CRP ≥ 0.2 mg/dL) with an adjustment for age and income to explore the possible role of Vit D in reducing complications attributed to unregulated inflammation and cytokine production.

**Data Sources:** Daily admission, recovery and deceased rate data for patients with COVID-19 were collected from Kaggle as of April 20, 2020. Screening data were collected from Our World in Data and official statements from public authorities. The mean concentration of 25(OH)D among the elderly for comparison with A-CMR was collected from previously published studies from different countries. Chronic factor data used in regression analysis was obtained from published articles. The correlation between Vit D and CRP was calculated based on 9,212 subject-level data from NHANES, 2009-2010.

**Results:** A link between 25(OH)D and A-CMR in the US, France, Iran and the UK (countries with similar screening status) may exist. We observed an inverse correlation (correlation coefficient ranging from −0.84 to −1) between high CRP and 25(OH)D. Age and the family income status also correlated to high CRP and subjects with higher age and lower family income presented more incidences of high CRP. Our analysis determined a possible link between high CRP and Vit D deficiency and calculated an OR of 1.8 with 95%CI (1.2 to 2.6) among the elderly (age ≥ 60 yo) in low-income families and an OR of 1.9 with 95%CI (1.4 to 2.7) among the elderly (age ≥ 60 yo) in high-income families. COVID-19 patient-level data shows a notable OR of 3.4 with 95%CI (2.15 to 5.4) for high CRP in severe COVID-19 patients.

**Conclusion:** Given that CRP is a surrogate marker for cytokine storm and is associated with Vit D deficiency, based on retrospective data and indirect evidence we see a possible role of Vit D in reducing complications attributed to unregulated inflammation and cytokine storm. Further research is needed to account for other factors through direct measurement of Vit D levels in COVID-19 patients.

## 1. Introduction

The recent global outbreak of COVID-19 imposed catastrophic impacts on every society, specifically among elderly populations. Currently, no treatment or vaccine against the virus is available. Consequently, there is a significant need to elucidate potential approaches that can reduce the number of severe COVID-19 cases and thus reduce the mortality rate associated with the disease. It has also been proposed that the immune system in some patients may manage COVID-19 better than in others. However, the potential causes or mechanisms underlying this have yet to be determined.

Temporal analysis of the number of confirmed, deceased and recovered cases across the world reveal patterns as to how COVID-19 has impacted different populations, which may help improve our understanding of the defense mechanism of the immune system against COVID-19 as well as aid in developing effective treatment options. Data show that the mortality rate of COVID-19 varies dramatically across countries. For example, a higher case fatality ratio has been reported in Spain, Italy, and the UK compared to that of the US and Germany. The cause for these disparities is not well understood. Several hypotheses have been proposed, including the emergence and circulation of different strains of the virus[1–3], idiosyncrasies in COVID-19 testing strategies and policies across countries, quality and access to health care, demographic factors such as the prevalence of elderly within a given population, and socioeconomic factors[4]. Some studies have suggested an analysis of age-specific case fatality ratio (CFR) and time-adjusted case mortality ratio (T-CMR) for a more insightful study of COVID-19 infection[5,6]. Initial reports and data obtained from various studies suggest that the elderly are disproportionately impacted by COVID-19[7]. The substantially higher CFR of the elderly population thus compels an age-specific analysis of COVID-19 data.

Aging can lead to a weakening of the innate immune system[8] which may play a role in the development of severe COVID-19. Specifically, a weak innate immune system response in the elderly can lead to a higher load of SARS-CoV-2 and a consequent overactivation of the adaptive immune system, leading to an increased level of cytokine production[9]. Clinical data obtained from COVID-19 patients in China showed high concentrations of cytokines such as granulocyte colony stimulating factor (GCSF), interferon gamma inducible protein 10 (IP10), macrophage chemotactic protein-1 (MCP1), macrophage inflammatory protein (MIP)1A, and tumor necrosis factor (TNF)α in patients admitted to the ICU, which suggests the presence of cytokine storm in these cases[10].

The role of Vit D in regulating the immune system has been supported by multiple studies[11]. Vit D can suppress cytokine production by simultaneously boosting the innate immune system (thus reducing the viral load) and decreasing the overactivation of the adaptive immune system to immediately respond to the viral load. Some researchers have suggested the potential role of Vit D in suppressing cytokine storm during the 1918-1919 viral influenza pandemic[12]. Moreover, the role of Vit D in enhancing immune response in flu and previous coronaviruses has been suggested[11,13]. It is this ability of Vit D in suppressing cytokine production[14,15]that motivated our focus on Vit D deficiency and its association with severe COVID-19.

To the best of our knowledge, no randomized blinded experiment has yet reported Vit D status and cytokine levels in patients with COVID-19. In spite of this, it is still possible to investigate the association between Vit D status and unregulated inflammation and cytokine production leading to severe COVID-19 based on a potential link between Vit D deficiency and C-reactive proteins (CRP)[16].

CRPs are produced primarily in the liver in response to inflammation to minimize damage to tissues from autoimmunity, infection, and other causes. The inflammatory cells’ ability to convert Vit D metabolites into calcitriol (the active form of Vit D) and to express the nuclear receptor of Vit D suggests a potential inverse association between CRP and Vit D, which is also supported by epidemiological studies[17,18]. Early studies have shown that calcitriol treatment attenuates both CRP and inflammatory cytokines (CD4(+) IFN-γ) in hemodialysis patients[19]. Researchers have proposed that calcitriol modulates cytokine levels (such as TNF-α and IL-1β) through the intercellular role of calcium[20,21].

Here we combine Vit D and CRP data from NHANES, 2009-2010 dataset[22] with clinical data from COVID-19 patients[23] to investigate the potential role of Vit D in regulating inflammation and cytokine production leading to severe COVID-19 across different countries. We partially address some of the concerns regarding association between Vit D and other risk factors of COVID-19 A-CMR including heart disease, diabetes, age, and obesity in each country via regression analysis.

## 2. Methods

Data regarding the number of affected cases, deaths, and recoveries from COVID-19 was obtained from Kaggle[24] as of April 20, 2020. Data regarding cases that have undergone testing were obtained from Our World in Data[25]. Age distribution of confirmed cases, those admitted to ICU, and deceased patients in Spain was based on data available from the Spanish Ministry of Health[26]. The concentration of 25-hydroxyvitamin D (25(OH)D) among the elderly population in each country was obtained from prior studies[27–32]. CRP data, Vit D, data and demographic variables of the subjects were pooled the cross-sectional data from 2009-2010 NHANES, conducted by the National Center for Health Statistics (NCHS), Centers for Disease Control and Prevention (CDC) [22]. Data regarding the risk factors including blood pressure[33], body to mass ratio[34], and diabetes[35], were obtained from published articles. Data on coronary heart disease (CHD) death rates across different countries was based on the calculation of World Life Expectancy on data reported by the World Health Organization (WHO) [36]. The link between high CRP and severe COVID-19 was examined based on data from a study investigating the characteristics of COVID-19 patients in China[23]. The T-CMR is defined as the estimated ratio of deceased patients on day N (D_N_) to confirmed patients on day N-8 (C_N-8_). Adaptive averaging of T-CMR (A-CMR) was calculated based on a weighted average technique as shown in Equation (1).

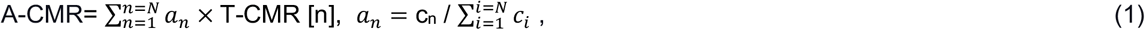

where N is the number of days with more than 10,000 confirmed cases in the country (except in S. Korea where the threshold is 5,000), c_i_ is the number of confirmed cases at day i, T-CMR (n) is T-CMR on day n, and a_n_ is a coefficient that describes the weight of T-CMR on day n. Positivity change (PC) is calculated using a moving average of size 5 on the ratio of daily confirmed cases to the daily tested individuals on day N as shown by Equation (2).

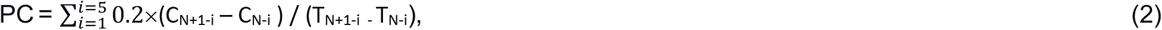

where C_N_ is the total confirmed cases on day N and T_N_ is the total number of tested cases on day N. Risks and conditional risks of the events were estimated using the ratio of the number of events in a treated group to the number of patients in the group. High CRP was defined as ≥0.2 mg/dL among healthy subjects (threshold suggesting low-grade inflammation and risk of cardiovascular diseases) and ≥1 mg/dL for COVID-19 patients.

## 3. Results and Interpretation

### 3.1. COVID-19 Fatality

Ambiguity in the incubation period of COVID-19 makes the calculation of the true mortality rate for the disease a challenging task[5,6]. Bureaucratic screening policies, as well as demographic and cultural variables further increase the difficulty of estimating disease onset and calculating an accurate case mortality rate (CMR). Analysis of time events reported from 41 deceased patients in Wuhan (Hubei, China) shows a median time of 8 days between admission and time of death, and 14 days between the onset of symptoms and time of death (shown in the inset in Figure 1 (a))[37]. This suggests a delay between the time the confirmed cases are reported and the time deceased patients are counted. In other words, the total number of deceased patients at day N (D_N_) is attributed to the total number of confirmed patients at day N-8 (C_N-8_) which is equal to the total number of cases at the onset of the symptoms on day N-14 (O_N-14_). Time adjusted-CMR (T-CMR) with a delay of 8 days (D_N_/ C_N-8_) is therefore used in this study (shown in Figure 1(a)). Calculating the percent difference between T-CMR on April 20 and April 6 for three different delays of 0 days, 8 days and 14 days suggests that an 8-day delay presents the least variation across countries.

**Figure 1.**
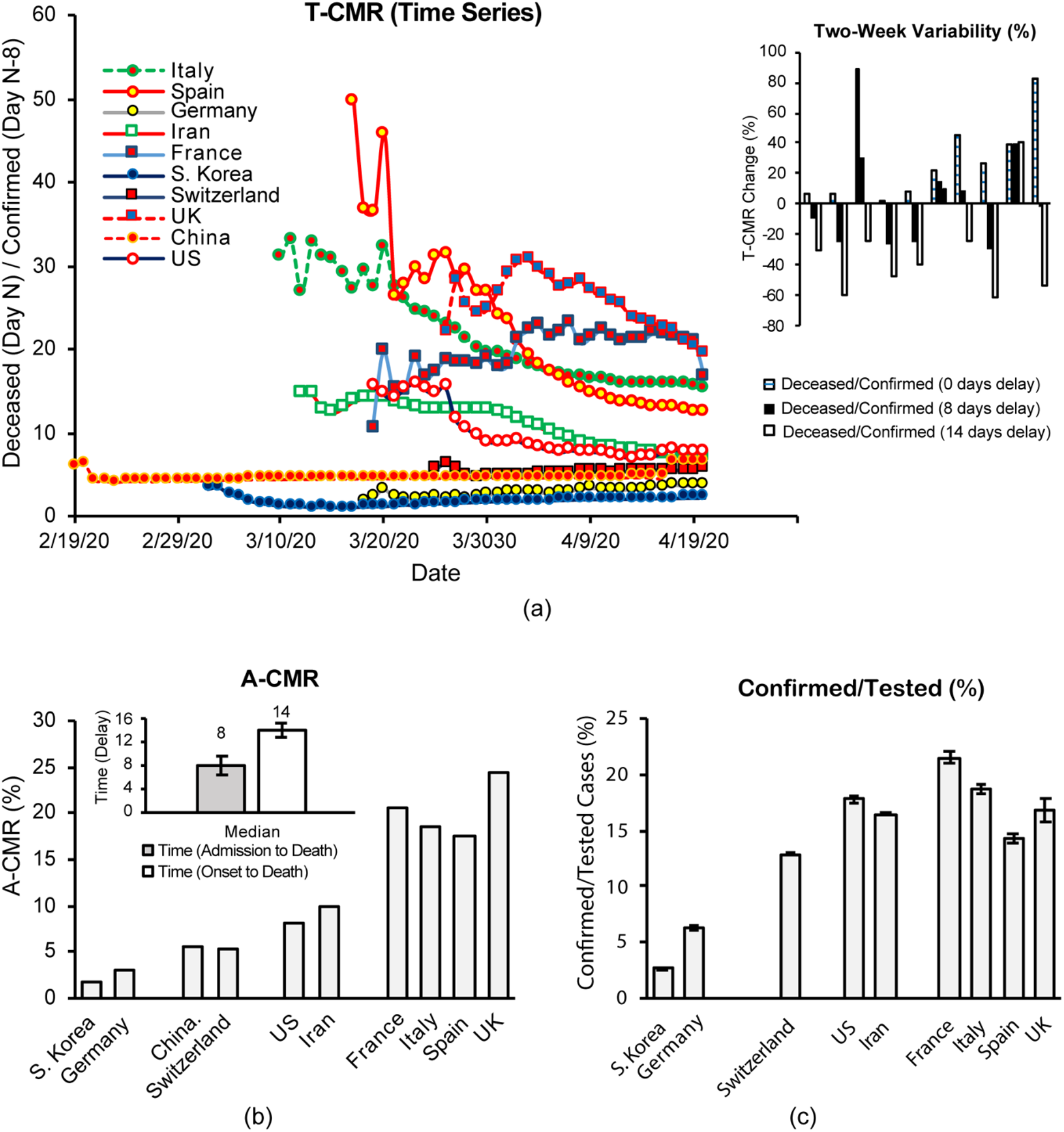
(a) The T-CMR (8 days) as of April 20. A two week variability (100 × (T-CMR_April 20_ − T-CMR_April 6_)/ T-CMR_April 6_) calculated at different T-CMR delays of 0 days, 8 days and 14 days. (b) A-CMR as of April 20 [24]. (c) Percentage of confirmed to tested ratio suggests an impact of screening policies in different countries on A-CMR[24,25,38–40]. France has reported the number of tests[25]. The UK data is reported number of tests (from April6-April 20) and we estimated number tests before April 6 by multiplying the number of tested patients by 1.26 (estimated from the relation between the number of tests and patients after April 6-April 20)[25]. US data is mainly the number of people tested (some labs have reported the number of tests)[25]. The number of tests conducted in Iran and Spain is estimated from two reported statements by public authorities[25,38,40].

This analysis acknowledges that the calculation of T-CMR with an 8-day delay is less sensitive to an abrupt change in the number of confirmed/deceased patients in a single day for a given country. Figure 1(a) shows time series data for T-CMR drifting for some countries. Intense variations in the ratio of confirmed to tested patients can change the results for T-CMR over the course of the pandemic for multiple reasons. With the deaths of the most vulnerable members of a population, T-CMR is expected to decrease over time. In addition, increasing screening capabilities will increase the chance of identifying mild cases, thus reducing T-CMR. As a result, different values for T-CMR are calculated throughout the pandemic and the question arises as to which value is more representative of the intrinsic mortality characteristic of the virus within each country.

#### A-CMR

In countries with a wide variation in confirmed cases, T-CMR varies each day and this increases the uncertainty of the actual T-CMR within the country. To calculate a more accurate estimate of T-CMR we created a framework based on two factors. First, we considered only outbreaks of 10,000 confirmed patients or greater (except in S. Korea where the threshold was set to 5,000 as the total confirmed cases stayed below 10,000 until April 3, 2020) to provide a reliable T-CMR. Next, an average of the T-CMRs was calculated given a higher weight for the T-CMRs that represent a higher population. A-CMR for each country is calculated using Equation (1) and the results (shown in Figure 1 (b)) suggest varying A-CMR values across countries.

S. Korea and Germany report a comparably low A-CMR of 1.8% and 3.1%, respectively. The A-CMR in Switzerland (A-CMR = 5.3%) and China (A-CMR = 5.5%) is higher than in S. Korea and Germany but is lower than in the US (A-CMR = 8%) and Iran (A-CMR of 9.8 %). Spain (A-CMR = 17.5%), Italy (A-CMR = 18.6%), France (A-CMR = 20.7%) and the UK (A-CMR = 24.5%) report the highest A-CMR. Multiple factors may contribute to the difference in A-CMR across these countries. Figure 1 (c) shows the average ratio of confirmed (C) to tested (T) cases in each country. Comparison of Figure 1(b) and 1(c) shows that countries with mass screening policies (low C/T ratio) report a substantially lower A-CMR than other countries. One reason could be that countries with an aggressive screening policy tend to detect more cases of mild, less deadly COVID-19 and will thus report a lower A-CMR, as mild COVID-19 cases are generally not fatal. In addition, a slow growth rate of confirmed patients may potentially cause a different and less fatal version of the virus to circulate across countries as the more fatal version of the virus is contained in hospitals. When the virus is spreading across the country with a fast growth rate, more fatal versions of the virus have more ways to spread, which may result in an increase in A-CMR for a given country. Therefore, we consider positivity (C/T) or PC to be a better indicator of the impact of screening policy than total tests per capita. The reason is that a low number of tests per capita can be used when the total number of patients is low, but as the number of patients increases substantially more tests per capita are required to facilitate detection of mild COVID-19 cases. Age distribution of the population is also another crucial factor that impacts mortality for a given country as COVID-19 has been shown to be deadlier in elderly patients[7]. In the following section, information obtained from PC and prevalence is used to conduct a more in-depth analysis of screening strategies in different countries.

#### Screening Status

It is important to control for screening strategies and age distribution across countries before comparing Vit D status, as such variables may notably impact A-CMR. Two factors can be used to evaluate the screening status in different countries; 1) PC, and 2) the prevalence of COVID-19. We first calculated the PC to provide an illustration of the variation in positivity in different countries over time in Figure 2. The average PC value in the first 14 days is calculated and the results are shown in the inset of Figure 2. Based on this analysis, we observed that S. Korea, Germany, and Switzerland have lower PC values, while Iran, the US, France, Italy, Spain, and the UK share higher PC values. The starting point of each curve is the day that the country reported at least 10,000 patients in total (except S. Korea > 5,000).

**Figure 2.**
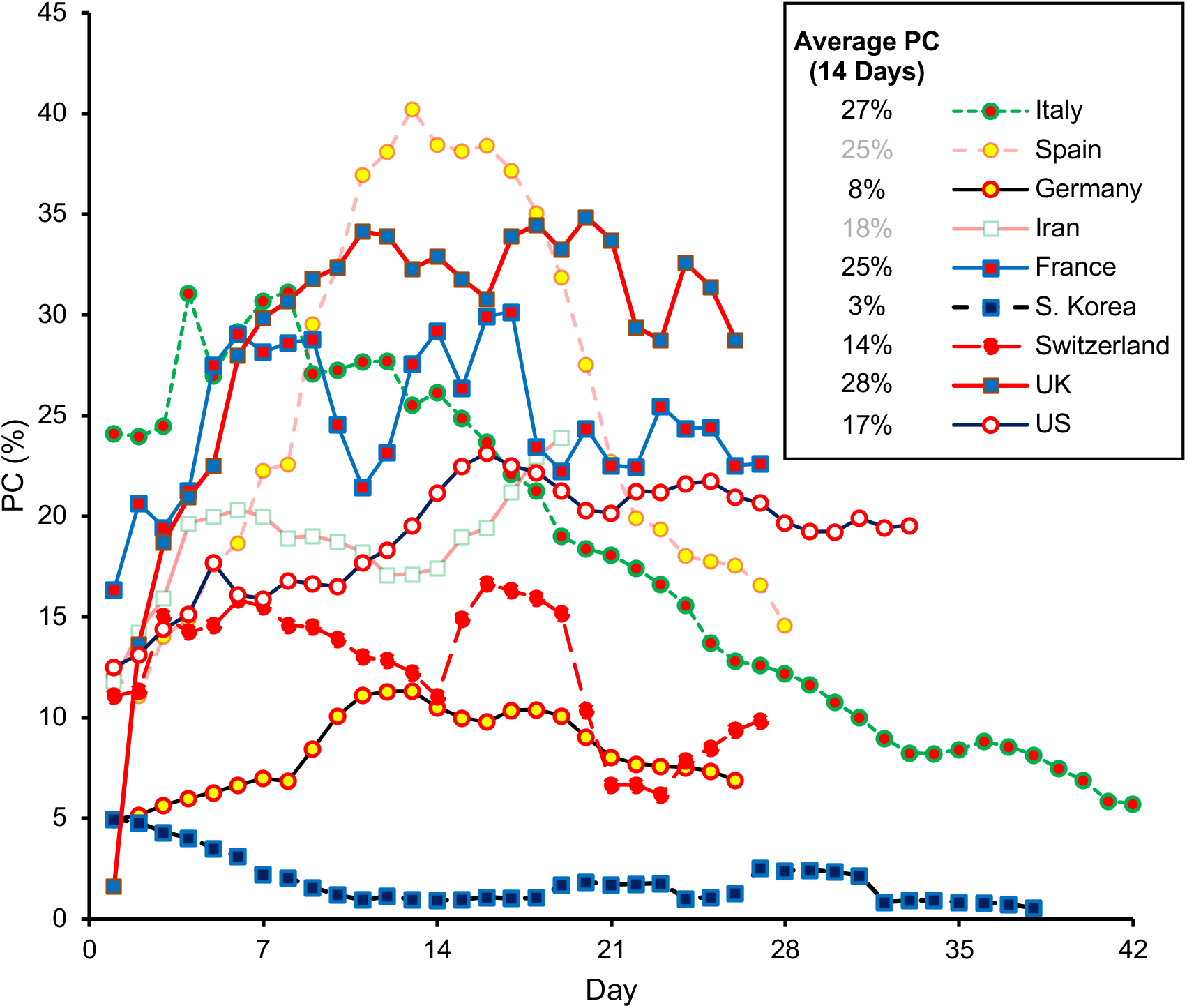
PC over time compares growth rate of COVID-19.

A weakness in this analysis is that positivity depends on the prevalence of COVID-19. Thus, we extended our analysis by evaluating PC as a function of prevalence. We calculated an average number of confirmed cases per 1 million population per day in 21 days (r_c_) and used it as an indicator of the prevalence of COVID-19 in each country. We plotted PC against r_c_ for two weeks in Figure 3 and the results suggest that the countries are clustered into two major groups where a more aggressive screening strategy is used such as in S. Korea, Germany and Switzerland compared to Spain, Italy, France, the UK, the US, and Iran. The initial state of each curve is similar to Figure 1 and Figure 2 where at least 10,000 confirmed patients (except 5,000 for S. Korea) are used to start the analysis. A testing aggressiveness index (TAI) is calculated using Equation (3) which presents a quantitative illustration for Figure 3.

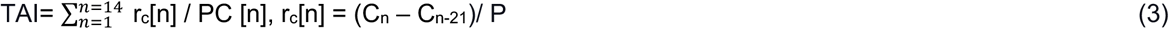

**Figure 3.**
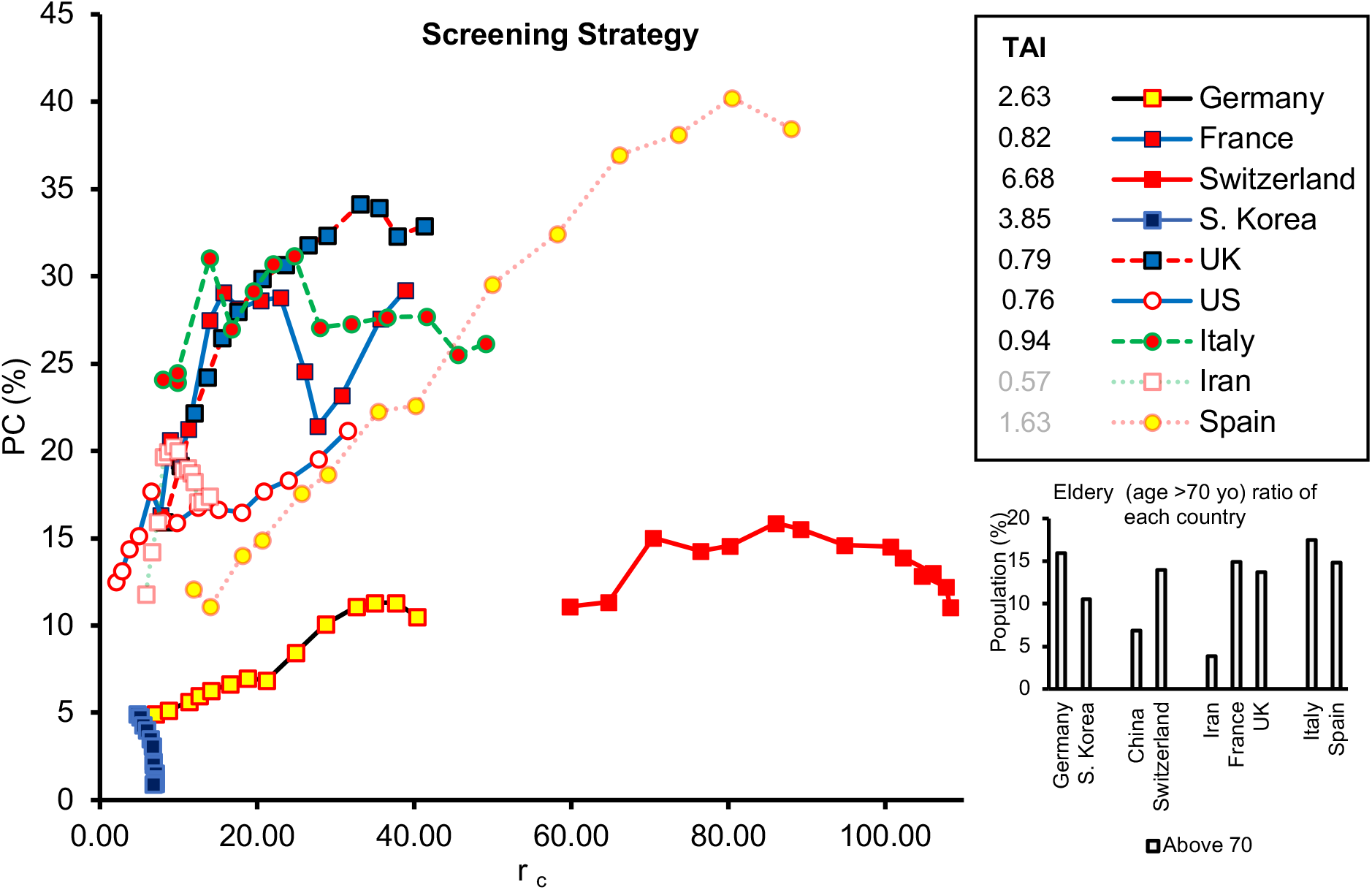
PC against r_c_ for two weeks after each country reaches 10,000 patients (except S. Korea >5,000 patients).

Where P is the population in millions of the countries, C_n_ is the total number of confirmed patients on day n. TAI values for each country are presented in the inset in Figure 3.

A small TAI is associated with a large delta in PC and a small delta in prevalence which indicates the population of the tested subjects (associated with PC) does not represent the number of confirmed subjects across the country’s population, thereby indicating a less aggressive screening strategy. The quantitative illustration of TIA suggests a more aggressive screening status (2.60<TIA<6.70) in Germany, S. Korea, and Switzerland, and a less aggressive screening status in Spain, Italy, France, the UK, the US, and Iran (0.55<TIA<1.65). The least aggressive screening status is found in Iran with TIA of 0.57. It should be noted that Spain and Iran have reported complete daily confirmed patient information but limited data about testing cases. The screening data from Iran and Spain are estimated from only two testing data points with an average new daily test rate reported by the public authorities. The limited number of data points may increase the error in our estimation, which is why these presented results are highlighted in gray in Figure 3. Furthermore, age distributions of different countries are shown in the inset of Figure 3 and suggest a similar age distribution between the US, the UK, France, Spain and Germany.

#### Possible Effect of Vit D on A-CMR

Screening status and the age distribution can notably impact A-CMR among the population of a given country. To evaluate the possible association of A-CMR with Vit D we need to ensure that both the screening status and age distribution of the elderly are similar between countries that are being compared.

#### Countries with Less Aggressive Screening Status

The 25(OH)D concentration among the elderly (age>60 yo or age>65 yo) in countries with less aggressive screening policies are shown in Figure 4 (a). A comparison of the A-CMR and the mean 25(OH)D concentration among the elderly suggests an inverse relationship between the two. In particular, the UK, with the lowest mean 25(OH)D level, reports the highest A-CMR while the US with the highest mean 25(OH)D reports the lowest A-CMR. Iran and France, countries with higher mean 25(OH)D concentration than the UK, report a lower A-CMR. The age distribution of the elderly among these countries, shown in the inset in Figure 3, indicates the US, France, and the UK have a similar elderly distribution while Iran and China have a lower elderly population than others.

**Figure 4.**
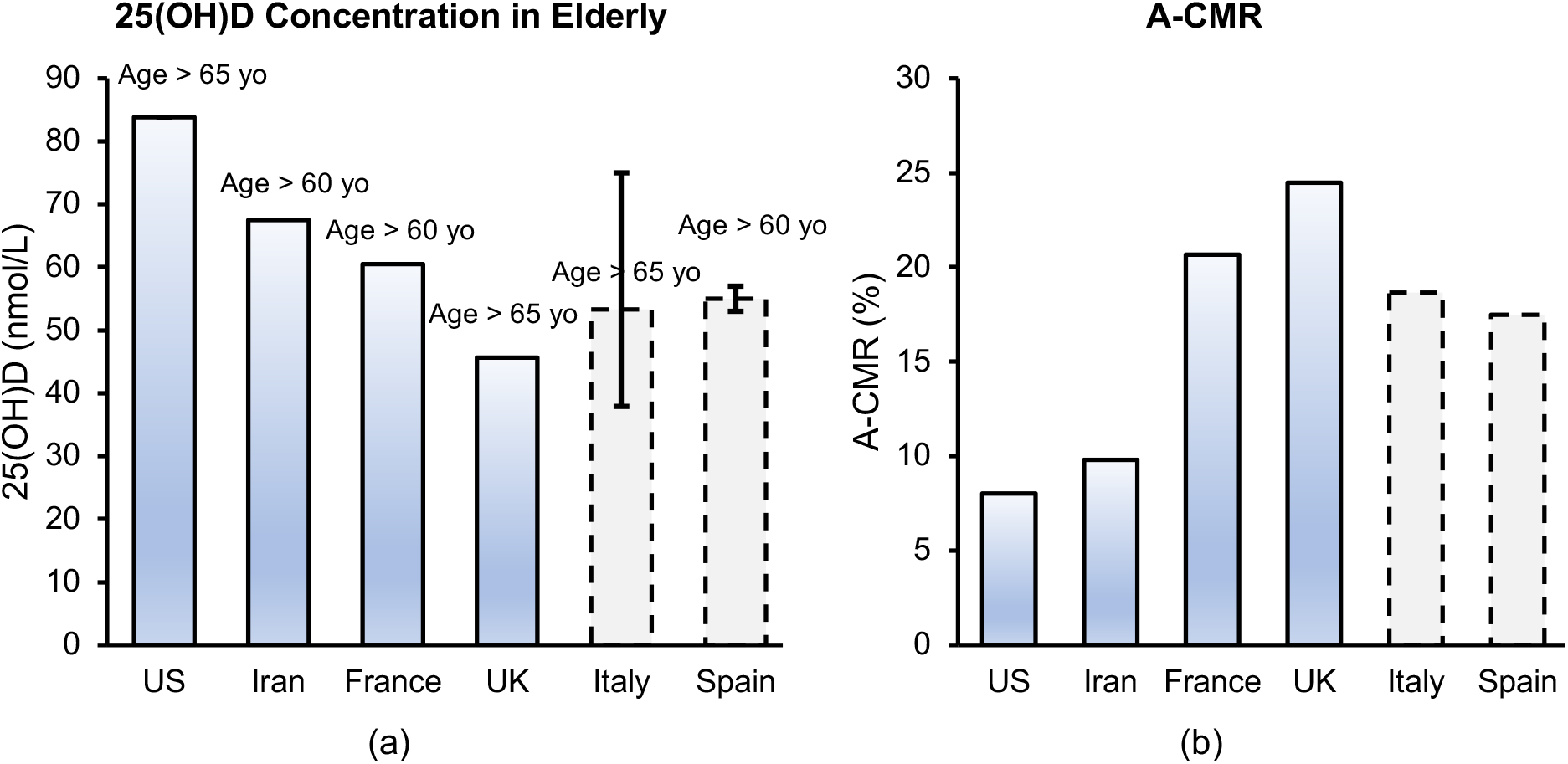
(a) Mean 25(OH)D in the elderly population in the US[41], Iran[42], France[43] and the UK[44], Average of three reported median 25(OH)D for Italy[27–29], Average of two reported median 25(OH)D among elderly Spain. Median of 25(OH)D in Spain has been estimated from Figure in the manuscript [30]. Error bar shows the range of reported median 25(OH)D in a different study, (b) A-CMR for the US, Iran, France, UK, Italy and Spain (countries with less aggressive screening status).

An estimate of 25(OH)D concentration from Italy and Spain is also included in this figure. Since we were unable to determine the mean 25(OH)D concentration for these countries, an average for the reported median of 25(OH)D concentration from different studies has been provided with a dashed bar. Studies involving different cohorts (the Asturias study and the Pizarra study) in Spain estimated a slightly different concentration of 25(OH)Damong the Spanish population. This led us to expect a median concentration between 53nmol/L to 59.5 nmol/L (values estimated from figure)[30]. The variation of reported 25(OH)D concentration of the elderly population in Italy was concerning. A study of 13,110 adults in Northwestern Italy estimated the median 25(OH)D concentration of 47 nmol/L among the elderly living there[27], while another study using data from 2,694 community-dwelling elderly from Northern Italy (results from the Progetto Veneto Anziani study) estimated a median of 75 nmol/L[28]. A third study of 697 elderly women in southern Italy estimated a mean 25(OH)D concentration of 37.9 nmol/L[29].

#### Countries with Aggressive Screening Status

Based on our analysis, Germany and S. Korea appear to have an aggressive screening strategy (Figures 2 & 3). The mean 25(OH)D concentration and A-CMR (shown in Figure 5) indicate that S. Korea is reporting a lower A-CMR than Germany while also reporting a higher mean 25(OH)D among the elderly. Although sensitivity analysis comparing Vit D status in the elderly population in countries with similar screening status suggests a possible role of Vit D deficiency in the elderly population affecting A-CMR, more countries need to be compared to better substantiate this analysis.

**Figure 5.**
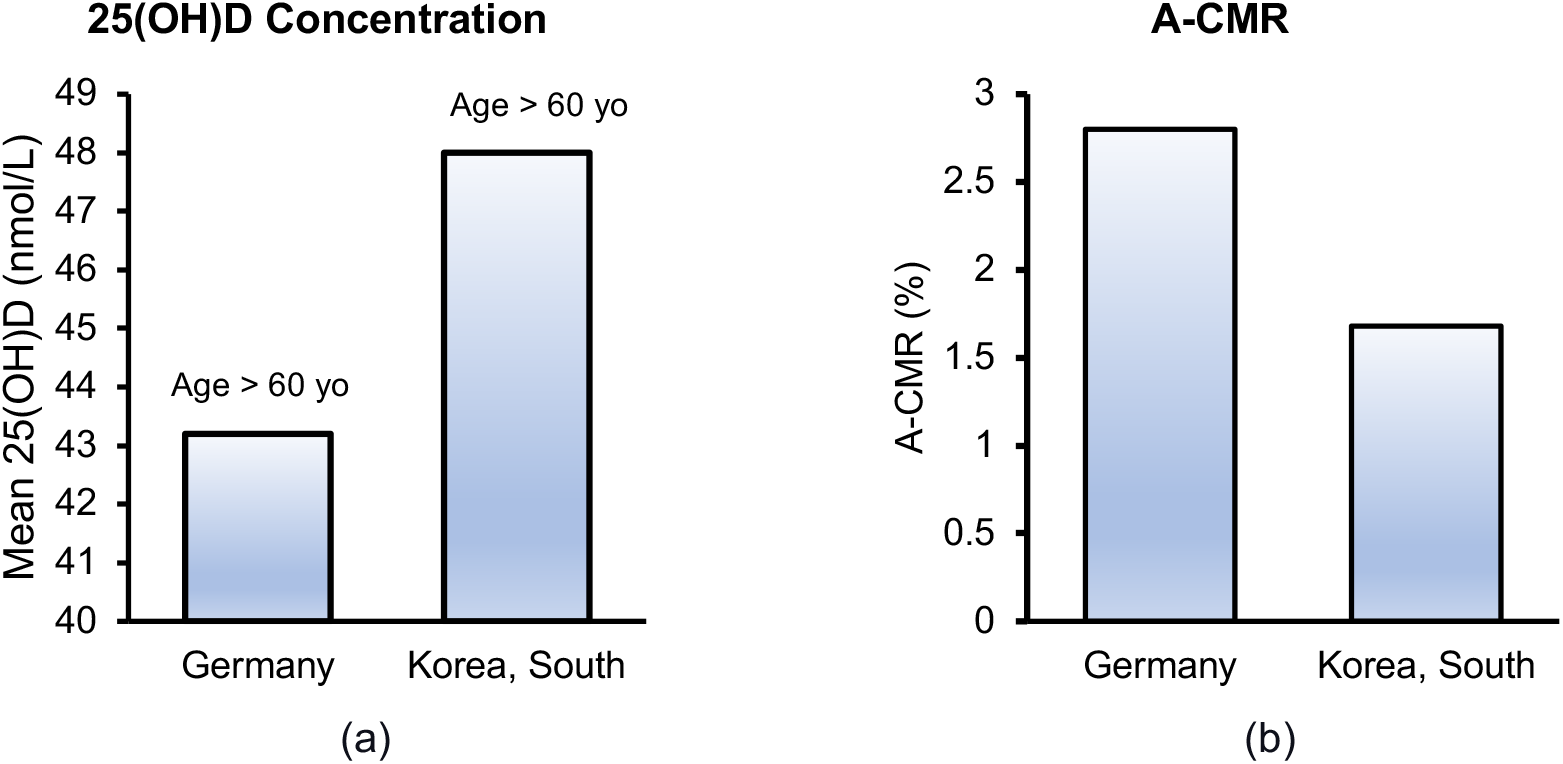
(a) Mean 25(OH)D concentration in the elderly population in Germany[31] and S. Korea[32]. (b) A-CMR in Germany and S. Korea.

#### Possible impact of Chronic Factors on A-CMR

The link reported between A-CMR and 25(OH)D can be impacted by chronic factors such as elderly ratio, heart disease prevalence, high blood pressure prevalence, body to mass ratio, and diabetes prevalence across the population. To investigate the possible impact of these chronic factors a regression model was made based on each variable and was used to predict the A-CMR among the countries with similar screening strategies. None of the investigated chronic factors were statistically significant and only 25(OH)D presented a p-value smaller than 0.05. Across these factors diabetes prevalence and CHD death per 100, 000 have a p-value between 0.1 and 0.15. A regression model based on only 25(OH)D (shown in Figure 6 (a)) can predict A-CMR with a root mean squared error of 3.1 while the regression model created based onthree chronic factors of diabetes prevalence (age-standardized), CHD death rate per 100, 000 (age-standardized), and elderly ratio (Figure 6(b)) can predict A-CMR with a root mean squared error of 7.1. This analysis partially addresses the limitation that our Vit D sensitivity analysis correlates with mortality due to its association with the underlying conditions such as diabetes, CHD, or age, however, we can not exclude residual confounding factors.

**Figure 6.**
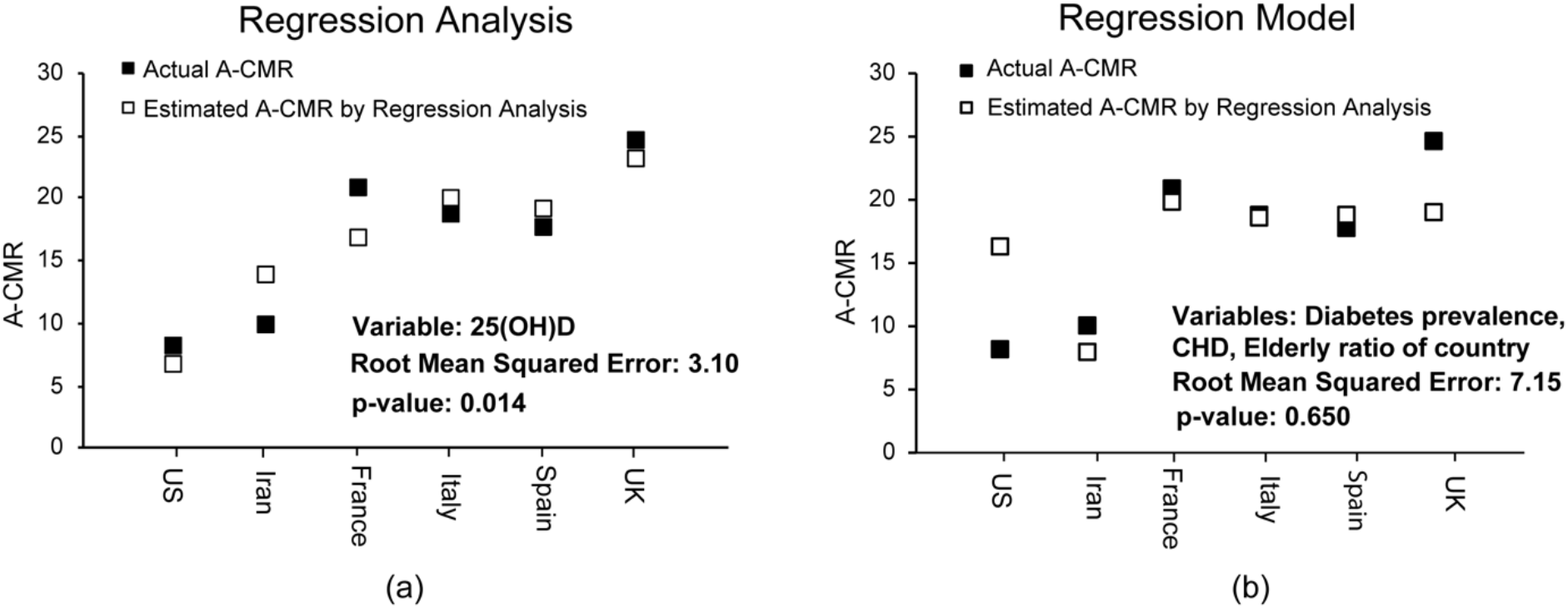
Regression analysis based on (a) 25(OH)D, (b) Diabetes prevelance among men and women (age standardized), elderly ratio (≥70 yo) in the country, CHD death rate per 100, 000 (age standardized)

### 3.2. Disparity in Confirmed, Hospitalized and Admitted to ICU Cases across Age Groups

The impact of aging on innate immunity may influence the body’s response against COVID-19. Figure 7 shows the age distributions of patients who were hospitalized, admitted to the ICU, and deceased based on 145,429 cases from Spain. It shows COVID-19’s alarming impact on the elderly. In particular, 61% of the patients above 70 yo were hospitalized and 20% died. Other studies have shown a similar vulnerability to COVID-19 among elderly populations[45–48]. The results shown in Figure 7 suggest a higher risk of hospitalization and admission to the ICU for elderly patients. A possible explanation for this is that a weak innate immune system response to COVID-19 can allow a high viral load, which then leads to a higher rate of complications and hospitalization. Consequent overactivation of the adaptive immune system and high levels of cytokine production[9] could then lead to complications that must be addressed in the ICU. A notably higher ratio of deceased to ICU-admitted patients among the elderly suggests that the impact of cytokine storm is more severe, and the threshold for tolerating its side effects is lower, for these patients. Figure 7 (b) also shows a higher ratio of admission to the ICU for children (<4 yo) which could be due to a relatively immature immune system[49]. A very low fatality rate among children suggests a higher threshold for tolerating complications associated with COVID-19 than among the elderly.

**Figure 7.**
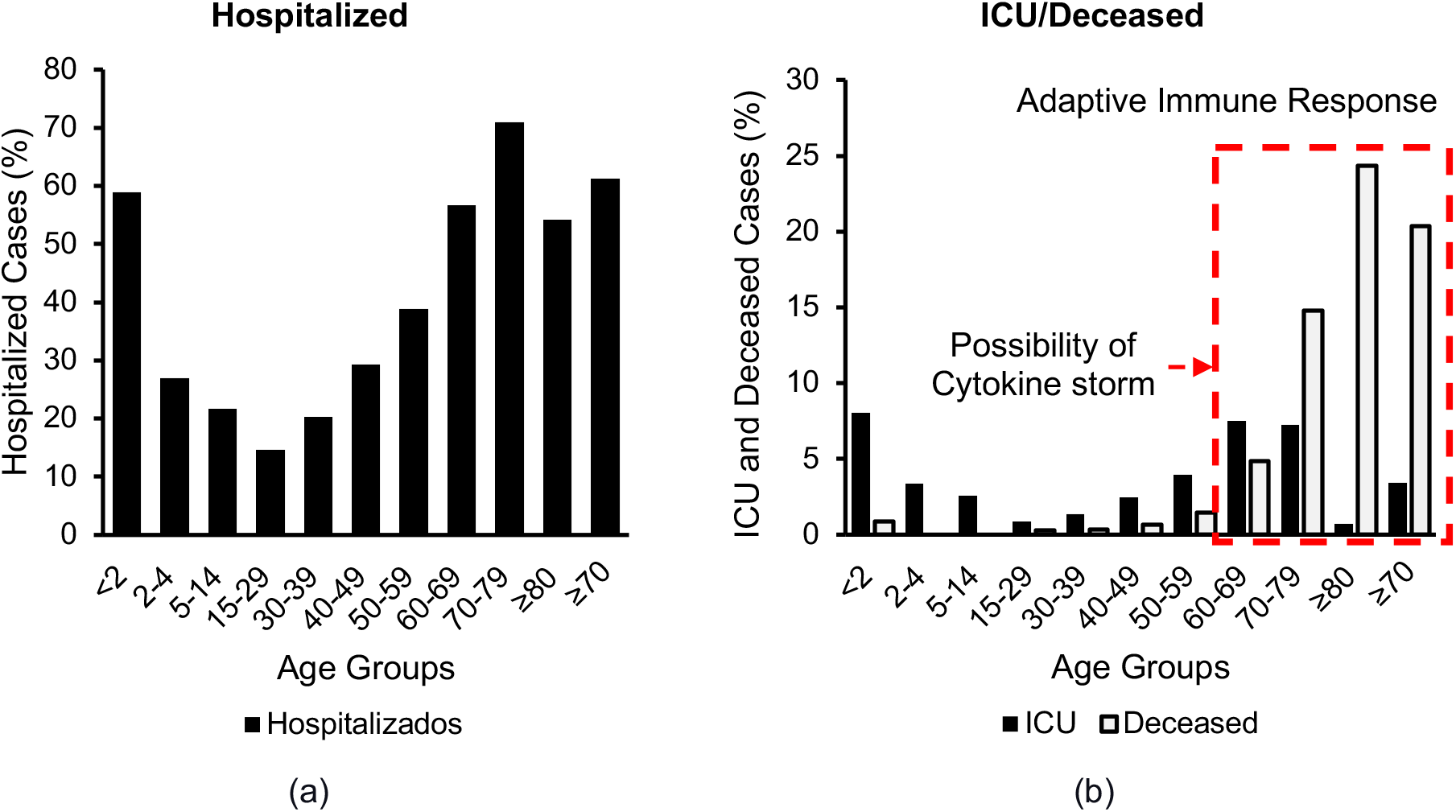
Age distribution of the a) hospitalized, b) admitted to ICU or deceased in Spain based on data from 145,429 cases[26].

### 3.3. CRP and Severe COVID-19

Table 1 shows the risks of severe and mild COVID-19 under different CRP levels, based on clinical data from 793 confirmed COVID-19 patients in China (up to 52 hospitals in 30 provinces)[23]. High CRP was defined as ≥1 mg/dL for COVID-19 patients associated with high-level inflammation and CRP produced via cytokine storm. According to this dataset, patients with severe COVID-19 have a higher incidence of high CRP (81.5%, 110 cases out of 135) than those with a mild form of the disease (56.5%, 371 cases out of 658). This suggests an Odds ratio (OR) of 3.4 with 95%CI (2.15 to 5.4). Conversely, patients with high CRP have a higher risk of severe COVID-19 (23%) than their counterparts with low CRP (8%). This trend also persists in cases of mild COVID-19.

**Table 1.** The risks of severe and mild COVID-19 under different CRP levels, based on data reported by[23].

|  | Number of Events/Total Patients (Risk) |
| --- | --- |
| **Risk of High CRP** | 481/793 (61%) |
| **Risk of Low CRP** | 312/793 (39%) |
| **Risk of High CRP given Severe COVID-19** | 110/135(81%) |
| **Risk of Low CRP given Sever COVID-19** | 25/135(19%) |
| **Risk of Severe COVID-19 given High CRP** | 110/481 (23%) |
| **Risk of Severe COVID-19 given Low CRP** | 25/312 (8%) |
| **Risk of High CRP given Mild COVID-19** | 371/658(56%) |
| **Risk of Low CRP given Mild COVID-19** | 287/658(44%) |

### 3.4. CRP and Vit D Deficiency in healthy Subjects

Studies have shown that Vit D deficiency leads to the production of cytokines such as TNF-α and IL-1p through the intercellular activity of calcium[20] which may lead to low grade inflammation followed by increased CRP levels. This may be the reason for the simultaneous attenuation of CRP and inflammatory cytokines (CD4(+) IFN-*γ*) in hemodialysis patients after calcitriol treatment[19], or elevation of both CRP and cytokines in severe COVID-19 patients[23]. The relationship between CRP and Vit D has been investigated in multiple clinical studies and demonstrating an inverse correlation between the two variables[16,50]. In the following we investigate the link between CRP and Vit D among different age groups and among subjects with low-income and high-income.

#### Association of Vit D Status with CRP at Different Age Groups

Our analysis of Vit D status and high CRP in similar age groups from 9211 participants (NHANES, 2009-2010), shown in Figure 8, indicates a higher incidence of high CRP among the elderly than the young subjects which could be due to more low-grade inflammation among the population.

**Figure 8.**
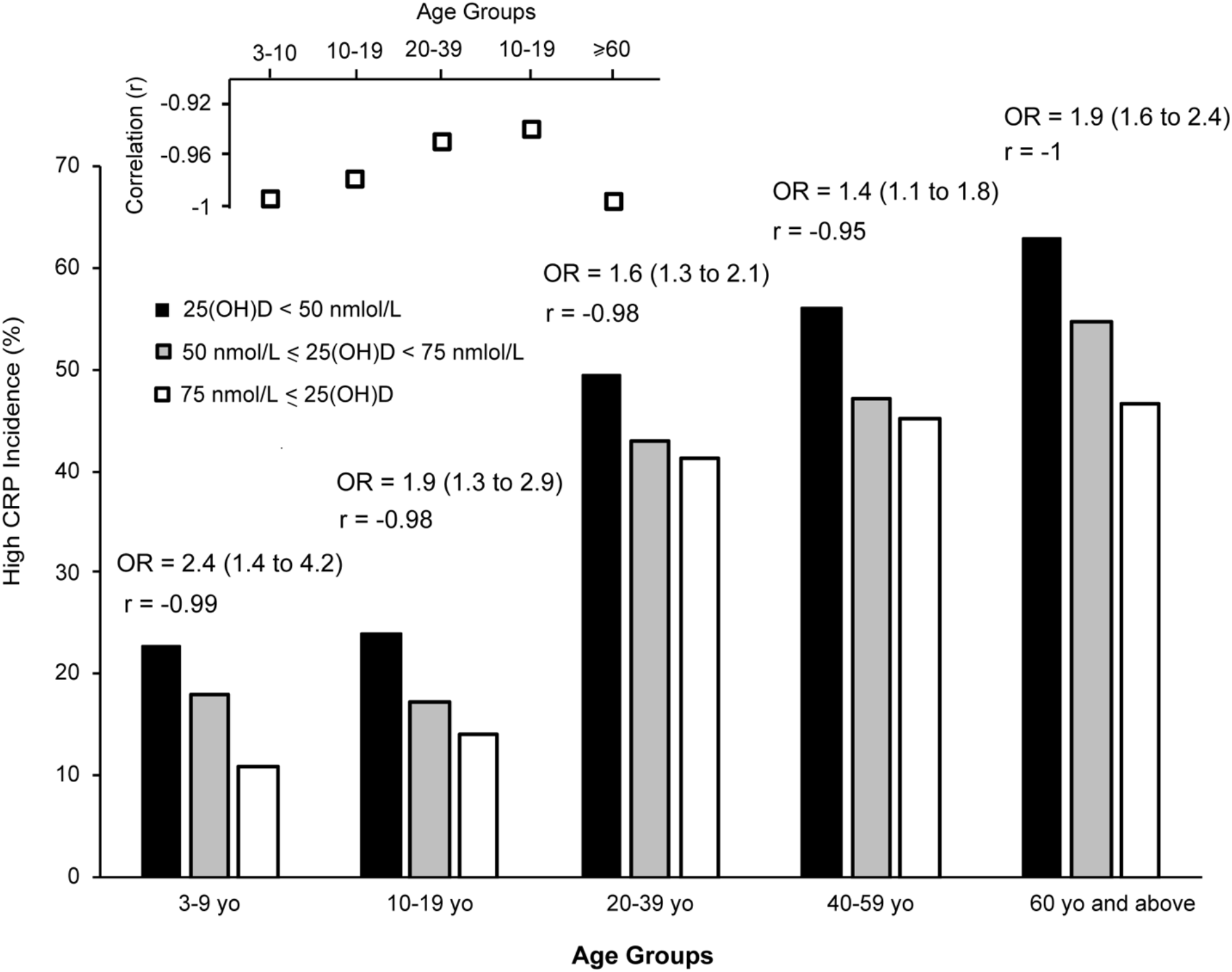
High CRP at different Vit D status in healthy subjects for different age groups

High CRP was defined as ≥0.2 mg/dL among healthy subjects which is the threshold suggesting low-grade inflammation and risk of cardiovascular disease[16]. Our analysis also shows subjects with Vit D deficiency are potentially producing more low-grade inflammation which causes higher incidents of high CRP than normal subjects. This is observed among all age groups. OR and correlation coefficients are calculated and shown above each bar. On average the incidence of high CRP is 12.1 (9.9 −14.2)% higher in subject with Vit D deficiency than the subjects with normal Vit D level. The correlation coefficient (r) ranged from −0.93 to −1.0 among different age groups suggests a strong inverse link between the two variables. Among the elderly above 60 yo, this analysis suggests an OR of 2.1 and correlation coefficient of −1.0 between CRP and Vit D for the elderly above 60 yo.

#### Association of Vit D Status with CRP at Different Age Groups with Similar Household Income (poverty baseline adjusted)

Investigating the link between the demographic variables and high CRP, we found a notable correlation between the ratio of family income to poverty and high CRP. This variable was calculated by dividing the total income of a family by a poverty index which was calculated based on guidelines described by the Department of Health and Human Services’ (HHS)), considering factors such as family size, state, and year. In our analysis Vit D and CRP data for 4526 subjects with a lower income to poverty index (0-2) are associated with low-income families while 3819 subjects with a higher Index (≥2) are associated with high-income families. To eliminate the potential impact of this variable from our analysis investigating the link between CRP and Vit D status, we determined the association between the Vit D status and high CRP at different age groups for both high-income and low-income and results for this analysis are shown in Figure 9. The odds ratio and r are presented next to each curve. This analysis shows that subjects with Vit D deficiency present a higher incidence of high CRP in all age groups among both low-income and high-income families than subjects with normal Vit D status. High CRP incidence in elderly ≥ 60 with Vit D deficiency is 15.5% (14.3% – 16.5%) higher than in elderly with normal Vit D status. This suggests elderly with Vit D deficiency have a notably higher risk of developing low-grade inflammations compared to the patients with normal Vit D status. Interestingly, subjects from low-income families have a higher risk of high CRP than the subjects from a high-income family which suggest they have a higher risk of low-grade inflammation than subject from high-income families. Considering CRP as a surrogate of cytokine production we predict a higher level of cytokines is possibly produced in elderly with Vit D deficiency.

**Figure 9.**
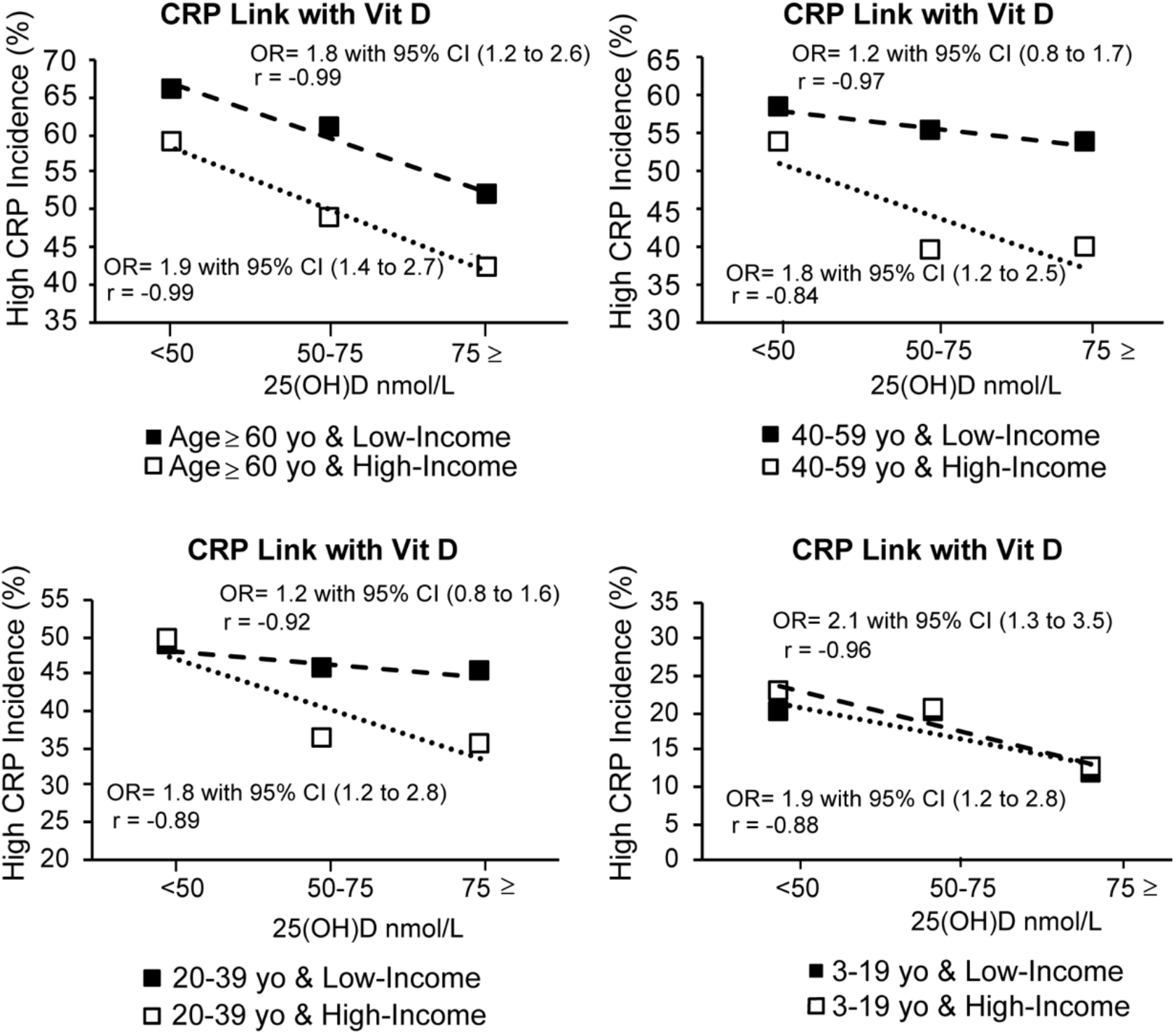
High CRP and possible low-grade inflammation association with Vit D status.

### 3.5. Possible Association between CRP, Cytokine Storm and Vit D deficiency

Production of IL-6 by monocyte, dendritic cells, and macrophage in patients with severe COVID-19 leads to systematic cytokine and CRP production[51]. CRP production will continue in response to the inflammation produced by the cytokine storm leading to high CRP levels. Patient-level data suggests higher cytokines and CRP levels in patients with severe COVID-19 than patients with mild COVID-19. Clinical data reported by Guan et al (summarized in table 2) indicates the risk of high CRP in severe COVID-19 patients is 44.5% higher than patients with mild COVID-19. Our analysis of CRP data of healthy subjects reported in Figure 9 after income adjustment suggests subjects with Vit D deficiency have 34% (age ≥ 60 yo), 22% (20 yo ≤ age <40 yo), and 21% (40 yo ≤ age <60 yo) more incidence of high CRP than patients with normal Vit D status. This may be due to the production of more low-grade inflammation in patients with Vit D deficiency than the patients with normal Vit D status and includes the inflammations caused by bioactivation of IL-6 and other cytokines. CRP is widely considered as a surrogate marker of IL-6 bioactivity[52–54] and the role of IL-6 in the development of cytokine storm in COVID-19 patients indicates the importance of CRP in the assessment of the related complications. Further studies using CRP, cytokine levels, and Vit D status from the same COVID patients before and after the infection is required to illustrate the impact of Vit D on the reduction of cytokine storm and severe COVID-19 complications. Our analysis reveals a possible role of Vit D in reducing cytokine levels and CRPs (attributed to unregulated inflammation) based on retrospective data and indirect evidence. Possible mechanism and pathway that Vit D can enhance regulating of cytokine levels are suggested in Figure 10.

**Figure 10.**
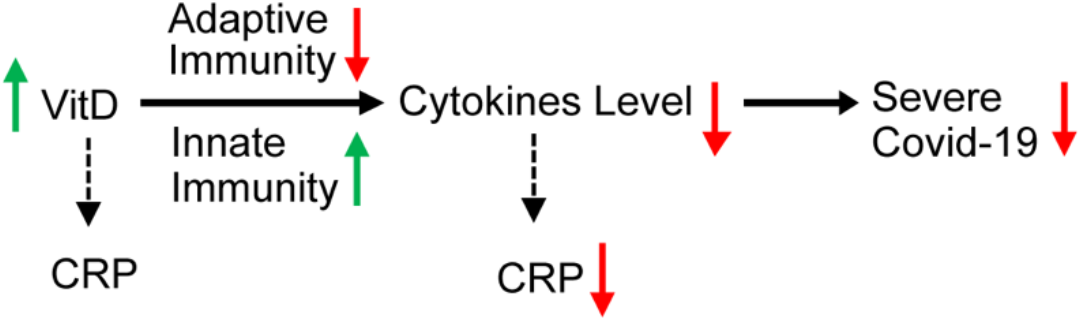
Possible impact of improving Vit D status in patients with severe Vit D deficiency on the reduction of cytokine levels.

While this epidemiological study provides compelling correlational evidence, we acknowledge that it clearly does not speak to causation. Indeed, while low Vit D levels have been associated with a variety of conditions (coronary artery disease, diabetes, cancers, autoimmune, obesity and other factors)[50], many of randomized controlled trials on Vit D supplementation have been disappointing[55,56]. This may be related to trial issues including the time frame of intervention, Vit D receptor polymorphisms, and the need to consider complementary or synergistic interventions, but underscores the need for caution. An alternative explanation for our findings may be that low Vit D status is a marker for underlying health issues which is known to be a risk factor in COVID-19 fatalities. Balancing this out is the strong biological/mechanistic plausibility for Vit D’s direct role in COVID-19 mitigation. This highlights the urgency for future randomized controlled trials.

## 4. Discussion

Our analysis of large-scale data suggests a possible link between Vit D deficiency and A-CMR among the countries with similar testing strategies. Another recent study comparing mortality in countries in the Southern and Northern Hemispheres, also supports the possible role of Vit D in severity of COVID-19[57].

We hypothesize that unregulated inflammation due to bioactivity of IL-6 and production of cytokine storm plays a role in COVID-19 complications affecting the A-CMR across different countries. Clinical data on cytokine levels in COVID-19 patients[58] points to the possible role played by the innate and adaptive immune systems in causing the elderly to be disproportionately hospitalized and admitted to ICU. Severe COVID-19 patients show a notable elevation of inflammatory cytokines such as interleukin (IL)-2R, IL-6, granulocyte colony-stimulating factor (GCSF), Macrophage chemotactic protein-1 (MCP1), macrophage inflammatory protein (MIP)1A, tumor necrosis factor (TNF)α and anti-inflammatory compounds such as CRP[10,58]. Complications associated with cytokine storm include Acute Respiratory Distress Syndrome (ARDS), exacerbation of the effects of pneumonia, acute kidney failure, acute heart failure, and rhabdomyolysis[24] which may become fatal. Elderly patients with an aberrant innate immune system may be subject to elevated viral load^4^ followed by misfiring and over-activation of their adaptive immune system through differentiating CD8+ T cells into Cytotoxic T Lymphocytes (CTLs)[59] potentially resulting in a cytokine storm. Of particular note is that the time interval for the development of a substantial adaptive immune response, approximately 7 days after development of symptomatic disease, is consistent with the time course of COVID-19 mortality[10,23]. The reported potential role of ibuprofen in worsening COVID-19 treatment[60] might also be partially explained by its suppression of innate immunity[61,62] which may lead to a higher viral load and consequent overactivation of the adaptive immune system which again may become fatal in elderly patients[63,64]. Even moderate lung damage due to a weak cytokine storm could lead to hypoxemia that in turn results in mortality due to underlying conditions.

Other studies have demonstrated the role of Vit D in regulating the immune system. Vit D may suppress cytokine production by simultaneously boosting the innate immune system and reducing the overactivation of the adaptive immune system in response to viral load[11,13]. Vit D deficiency may be more prevalent among the elderly and African-American[8] but it includes every population group.

One important limitation of the present country-level analysis is the assumption that Vit D levels in COVID-19 patients follow the same distribution with subjects in other previous Vit D studies. We did not have access to Vit D status and cytokine levels in individual COVID-19 patients. In other words, we do not have the data to suggest that Vit D is therapeutic. Leveraging available data, we illustrated a possible association between Vit D and severe COVID-19 based on a potential link between Vit D deficiency and high CRP (a surrogate of cytokine storm). In addition, the difference in age range, ethnicity, gender, social status, geographic latitude, measurement variations, the season of sample collection, and year of study may impact the reported value of Vit D status in different studies. We have partially addressed the A-CMR correlation with the underlying conditions such as diabetes, CHD, or age in our regression analysis, however, we can not exclude residual confounding factors. Analysis of CRP, and cytokine levels, and Vit D status from the same COVID-19 patients before and after infection is required to show the impact of Vit D on COVID-19 patients. The intrinsic cross-sectional nature of this study does not prove a relationship between Vit D, CRP levels, cytokine storm, and severe COVID-19. Vit D data have been collected from different sources and variation between and within different studies introduces variations in the data. Another important limitation of this study is that crude mortality data is used instead of age-specific mortality data. This is because the age distribution of confirmed patients in the US and UK were not available. The onset of COVID-19 for confirmed cases is unknown and is assumed to be similar for all subjects. In addition, other underlying conditions associated with the populations at risk of Vit D deficiency makes it more challenging to assess the actual impact of Vit D in comparison to other factors. Finally, our data suggest differences in the COVID-19 testing strategies and policies across countries. This makes an accurate assessment of mortality difficult. To address this issue, in our analysis we grouped countries based on the presumptive similarities between their testing strategies. These limitations can be addressed by following Vit D and COVID-19 status in individual patients within a given population. Such data, however, is currently unavailable. The link between Vit D and the probability of severe COVID-19 and associated mortality that is indicated by this work may serve as an impetus for such studies.

## 5. Conclusion

Large-scale data shows that different countries have differing A-CMR among confirmed cases. Screening strategies notably impact A-CMR, as countries with aggressive COVID-19 screening show decreased A-CMR. Our analysis of mean 25(OH)D across countries with similar testing strategies determined a possible link between Vit D status and A-CMR. Our regression analysis shows other chronic factors such as the elderly ratio of population, the prevalence of diabetes, prevalence of CHD in a country have less impact on A-CMR than Vit D deficiency. Our hypothesis on the role of unregulated inflammation in COVID-19 complications, is consistent with findings such as an increase in the rate of complications with age, low rate of complications in children, and adverse outcomes with ibuprofen, and suggests that it might be of interest to study Vit D’s role in COVID in controlled observational or clinical trials. Our analysis of high CRP in healthy subjects (CRP ≥ 0.2 mg/dL) showed an inverse relation between Vit D status and high CRP which is an indicator of low-grade inflammation in healthy subjects. We showed elderly and subjects from low-income families are at a higher risk of the low-grade inflammations attributed to high CRP. We calculated an OR of 1.8 with 95%CI (1.2 to 2.6) among the elderly (age ≥ 60 yo) in low-income families and an OR of 1.9 with 95%CI (1.4 to 2.7) among the elderly (age ≥ 60 yo) in high-income families. COVID-19 patient-level data shows a notable OR of 3.4 with 95%CI (2.15 to 5.4) for high CRP in severe COVID-19 patients (CRP ≥ 1 mg/dL). Based on retrospective data and indirect evidence we see a possible role of Vit D in reducing complications attributed to unregulated inflammation and cytokine storm however we emphasize that we do not have the patient-level data to suggest that Vit D is therapeutic.

## Data Availability

Data regarding the number of affected cases, deaths, and recoveries from COVID-19 was obtained from Kaggle[24] as of April 20, 2020. Data regarding cases that have undergone testing were obtained from Our World in Data[25]. Age distribution of confirmed cases, those admitted to ICU, and deceased patients in Spain was based on data available from the Spanish Ministry of Health[26]. The concentration of 25(OH)D among the elderly population in each country was obtained from prior studies[27-32]. CRP data, Vit D, data and demographic variables of the subjects were pooled the cross-sectional data from 2009-2010 NHANES, conducted by the National Center for Health Statistics (NCHS), Centers for Disease Control and Prevention (CDC) [22]. Data regarding the risk factors including blood pressure[33], body to mass ratio[34], and diabetes[35], were obtained from published articles. Data on coronary heart disease (CHD) death rates across different countries was used based on the calculation of World Life Expectancy on data reported by the World Health Organization (WHO) [36]. The link between high CRP and severe COVID-19 was examined based on data from a study investigating the characteristics of COVID-19 patients in China[23].
References:
22 NHANES 2009-2010 Laboratory Data. https://wwwn.cdc.gov/nchs/nhanes/search/datapage.aspx?Component=Laboratory&CycleBeginYear=2009 (accessed 13 May 2020).
23 Guan W, Ni Z, Hu Y, et al. Clinical Characteristics of Coronavirus Disease 2019 in China. N Engl J Med 2020;0:null. doi:10.1056/NEJMoa2002032
24 Novel Corona Virus 2019 Dataset. https://kaggle.com/sudalairajkumar/novel-corona-virus-2019-dataset (accessed 1 Apr 2020).
25 To understand the global pandemic, we need global testing - the Our World in Data COVID-19 Testing dataset. Our World Data. https://ourworldindata.org/covid-testing (accessed 13 Apr 2020).
26 Informes COVID-19. https://www.isciii.es/QueHacemos/Servicios/VigilanciaSaludPublicaRENAVE/EnfermedadesTransmisibles/Paginas/InformesCOVID-19.aspx (accessed 4 Apr 2020).
27 Basile M, Ciardi L, Crespi I, et al. Assessing serum concentrations of 25-hydroxy-vitamin d in north-western Italy. J Frailty Aging 2013;2:174-8. doi:10.14283/jfa.2013.25
28 Toffanello ED, Perissinotto E, Sergi G, et al. Vitamin D and Physical Performance in Elderly Subjects: The Pro.V.A Study. PLOS ONE 2012;7:e34950. doi:10.1371/journal.pone.0034950
29 Adami S, Viapiana O, Gatti D, et al. Relationship between serum parathyroid hormone, vitamin D sufficiency, age, and calcium intake. Bone 2008;42:267-70. doi:10.1016/j.bone.2007.10.003
30 González-Molero I, Morcillo S, Valdés S, et al. Vitamin D deficiency in Spain: a population-based cohort study. Eur J Clin Nutr 2011;65:321-8. doi:10.1038/ejcn.2010.265
31 Rabenberg M, Scheidt-Nave C, Busch MA, et al. Vitamin D status among adults in Germany - results from the German Health Interview and Examination Survey for Adults (DEGS1). BMC Public Health 2015;15. doi:10.1186/s12889-015-2016-7
32 Park J-H, Hong IY, Chung JW, et al. Vitamin D status in South Korean population. Medicine (Baltimore) 2018;97. doi:10.1097/MD.0000000000011032
33 Zhou B, Bentham J, Cesare MD, et al. Worldwide trends in blood pressure from 1975 to 2015: a pooled analysis of 1479 population-based measurement studies with 19·1 million participants. The Lancet 2017;389:37-55. doi:10.1016/S0140-6736(16)31919-5
34 Abarca-Gómez L, Abdeen ZA, Hamid ZA, et al. Worldwide trends in body-mass index, underweight, overweight, and obesity from 1975 to 2016: a pooled analysis of 2416 population-based measurement studies in 128·9 million children, adolescents, and adults. The Lancet 2017;390:2627-42. doi:10.1016/S0140-6736(17)32129-3
35 Zhou B, Lu Y, Hajifathalian K, et al. Worldwide trends in diabetes since 1980: a pooled analysis of 751 population-based studies with 4·4 million participants. The Lancet 2016;387:1513-30. doi:10.1016/S0140-6736(16)00618-8
36 Coronary heart disease death rate by country. World Life Expect. https://www.worldlifeexpectancy.com/cause-of-death/coronary-heart-disease/by-country/ (accessed 13 May 2020).

https://www.kaggle.com/sudalairajkumar/novel-corona-virus-2019-dataset

https://population.un.org/wpp/Download/Standard/Population/

https://www.who.int/vmnis/en/

https://www.isciii.es/QueHacemos/Servicios/VigilanciaSaludPublicaRENAVE/EnfermedadesTransmisibles/Paginas/InformesCOVID-19.aspx

http://ncdrisc.org/data-downloads.html

https://www.santepubliquefrance.fr/maladies-et-traumatismes/maladies-et-infections-respiratoires/infection-a-coronavirus/documents/bulletin-national/covid-19-point-epidemiologique-du-15-mars-2020

https://www.epicentro.iss.it/en/coronavirus/

https://wwwn.cdc.gov/nchs/nhanes/Search/DataPage.aspx?Component=Laboratory&CycleBeginYear=2009

https://assets.publishing.service.gov.uk/government/uploads/system/uploads/attachment_data/file/551352/NDNS_Y5_6_UK_Main_Text.pdf

https://ourworldindata.org/covid-testing

## Acknowledgment

The authors would like to thank Benjamin D Keane for his assistance in preparing the manuscript. The authors would also like to acknowledge generous support from the Carinato Charitable Foundation, Mark and Ingeborg Holliday, Kristin Hudson & Rob Goldman, and Ms. Susan Brice & Mr. Jordi Esteve.

